# A survey of International Health Regulations National Focal Points experiences in carrying out their functions

**DOI:** 10.1101/2021.02.01.21250930

**Authors:** Corinne Packer, Sam F. Halabi, Helge Hollmeyer, Salima S. Mithani, Lindsay Wilson, Arne Ruckert, Ronald Labonté, David P. Fidler, Lawrence O. Gostin, Kumanan Wilson

## Abstract

**Background:** The 2005 International Health Regulations (IHR (2005)) require States Parties to establish National Focal Points (NFPs) responsible for notifying the World Health Organization (WHO) of potential events that might constitute public health emergencies of international concern (PHEICs), such as outbreaks of novel infectious diseases. Given the critical role of NFPs in the global surveillance and response system supported by the IHR, we sought to assess their experiences in carrying out their functions.

**Methods:** In collaboration with WHO officials, we administered a voluntary online survey to all 196 States Parties to the IHR (2005) in Africa, Asia, Europe, and South and North America, from October to November 2019. The survey was available in six languages via a secure internet-based system.

**Results:** In total, 121 NFP representatives answered the 56-question survey; 105 in full, and an additional 16 in part, resulting in a response rate of 62% (121 responses to 196 invitations to participate). The majority of NFPs knew how to notify the WHO of a potential PHEIC, and believed they have the content expertise to carry out their functions. Respondents found training workshops organized by WHO Regional Offices helpful on how to report PHEICs. NFPs experienced challenges in four critical areas: 1) insufficient intersectoral collaboration within their countries, including limited access to, or a lack of cooperation from, key relevant ministries; 2) inadequate communications, such as deficient information technology systems in place to carry out their functions in a timely fashion; 3) lack of authority to report potential PHEICs; and 4) inadequacies in some resources made available by the WHO, including a key tool – the NFP Guide. Finally, many NFP representatives expressed concern about how WHO uses the information they receive from NFPs.

**Conclusion:** Our study, conducted just prior to the COVID-19 pandemic, illustrates key challenges experienced by NFPs that can affect States Parties and WHO performance when outbreaks occur. In order for NFPs to be able to rapidly and successfully communicate potential PHEICs such as COVID-19 in the future, continued measures need to be taken by both WHO and States Parties to ensure NFPs have the necessary authority, capacity, training, and resources to effectively carry out their functions as described in the IHR.

## BACKGROUND

States Parties to the International Health Regulations (IHR) (2005) seek to “prevent, protect against, control and provide a public health response to the international spread of disease in ways that are commensurate with and restricted to public health risks, and which avoid unnecessary interference with international traffic and trade” (1). To date, 196 countries have agreed to be bound by them. As a legal framework, the IHR establish a global structure aimed at providing the world with improved public health security with the least possible interference in international traffic, trade, and human rights (2,3).

Under Article 4 of the Regulations, all States Parties must establish or designate National Focal Points (NFPs). NFPs must be national centres (not individual persons) capable of conducting the communications required under the IHR within their States and internationally (3).

According to Article 4.2, NFPs are responsible for

> (*a*) *sending to WHO IHR Contact Points, on behalf of the State Party concerned, urgent communications concerning the implementation of these Regulations, in particular under Articles 6 to 12; and* (*b*) *disseminating information to, and consolidating input from, relevant sectors of the administration of the State Party concerned…*.

Furthermore,

> *Each State Party shall notify WHO, by the most efficient means of communication available, by way of the National IHR Focal Point, and within 24 hours of assessment of public health information, of all events which may constitute a public health emergency of international concern within its territory in accordance with the decision instrument*…
>
> (Article 6)

The functions of the NFPs under the IHR mean that States Parties must provide NFPs with the authority, capacity, training, and resources to fulfil their responsibilities effectively.

The IHR Review Committee on the Role of the IHR in the Ebola Outbreak and Response found that many NFPs lack the authority, capacity, training and resources to effectively carry out their mandate regarding urgent, event-based communications as stated in Article 4 of the IHR(4). This third Review Committee also observed that there was limited knowledge by high level-officials of the role of NFPs. Already the Review Committee on Second Extensions for establishing national public health capacities and on IHR implementation concluded that one of the key impediments to IHR implementation remains the insufficient levels of capacity and authority of NFPs(5). Similarly, the first IHR Review Committee noted that “there are indications that NFPs are not yet a timely source of initial, early information on events” and that “notification appears to have a high threshold in some countries”(6). Despite the widespread establishment of NFPs and States Parties’ acceptance of the IHR, results of studies suggest technical and political barriers to the notification of events to WHO(2,7).

A study was commissioned by the World Health Organization (WHO) to further assess the experiences of NFPs in carrying out their functions under the IHR, identify good practices among NFPs, and describe challenges that NFPs report in implementing the IHR. This study, carried out by the authors of this article, consisted of both interviews and a survey completed by NFP representatives. The authors submitted a report on the findings from both the interviews and the survey to the WHO in 2020. This article summarizes the results of the survey portion of the study.

## METHODS

Our study objectives were to evaluate NFPs’ self-reported ability to perform their mandated functions and determine difficulties they face through semi-structured interviews and a structured survey. The study received ethics clearance from the University of Ottawa Research Ethics Board and the Ottawa Health Science Network Research Ethics Board.

We developed an online survey to collect the data needed for this study (see Appendix 1). The survey sought information across a broad range of quantifiable indicators regarding NFPs’ perceived ability to implement their functions under the IHR and their needs for additional support and guidance from WHO. We had conducted qualitative interviews (n=25) shortly before launching the survey. Themes that emerged from the interviews were added to the survey questionnaire.

In order to facilitate quantitative analysis of NFP input, the survey posed questions as dichotomous (Yes/No) questions or using a Likert scale. We shared the questionnaire with WHO officials in Geneva, and subsequently WHO Regional Offices in order to solicit feedback. The comments and suggestions from the WHO were incorporated into the final version of the survey. WHO in Geneva provided translation services for the questionnaire.

An email was sent to all 196 NFPs by WHO Regional Offices informing them that they would receive an invitation from our team to participate in the survey. The invitation letter from our team made the online questionnaire available to all NFPs. The questionnaire was provided to the representatives of all NFPs in their preferred language of communication based on WHO’s six official languages (English, Arabic, French, Spanish, Russian, and Chinese) via a secure internet-based system. Participants with limited internet access were provided electronic attachments of the questionnaire to download, complete, and return by email. Only three NFPs used the electronic attachment option.

Our letter of invitation indicated that one staff member from each NFP (chosen by the NFPs themselves) was to complete the survey on behalf of the NFP, but the survey could also be completed as a group if NFP staff members felt that this would more accurately convey their perspectives. Each NFP could submit only one completed survey. In order to promote participation, our team sent multiple reminder emails to NFPs that had not responded to the invitation or had not completed the questionnaire in full.

Eight States Parties submitted two surveys. We resolved the issue of duplicate submissions by including only the first completed survey in the dataset. Participants did not contact our team for any clarification of questions.

The questionnaire included nine open-ended questions aimed at obtaining deeper insights into specific issues, such as feedback on WHO guidance documents and tools. The questionnaire also gave participants the opportunity to share any information materials that they had produced that could serve as templates or guidance documents for other NFPs.

We generated summarized data for the completed questionnaires for each language through the Survey Monkey tool in a pdf format. Our team entered the responses to each question in a Microsoft Excel spreadsheet, combining the datasets from each language. The responses to each dichotomous question were tabulated, and figures were produced. The results for the questions that allowed respondents to enter narrative information were also gathered in Excel along thematic groupings. Narrative responses provided in French and Spanish languages were translated into English by our team while Google Translate was used to translate responses in the other languages.

## RESULTS

Out of 196 IHR State Parties, 105 completed the questionnaire in full (53.6%), while 16 (8.2%) submitted partially-completed questionnaires. Nearly half the submissions came from two regions, Europe and Africa (Figure 1). The most frequent language selected was English, followed by French, Spanish, Russian, and Arabic.

**Figure 1:**
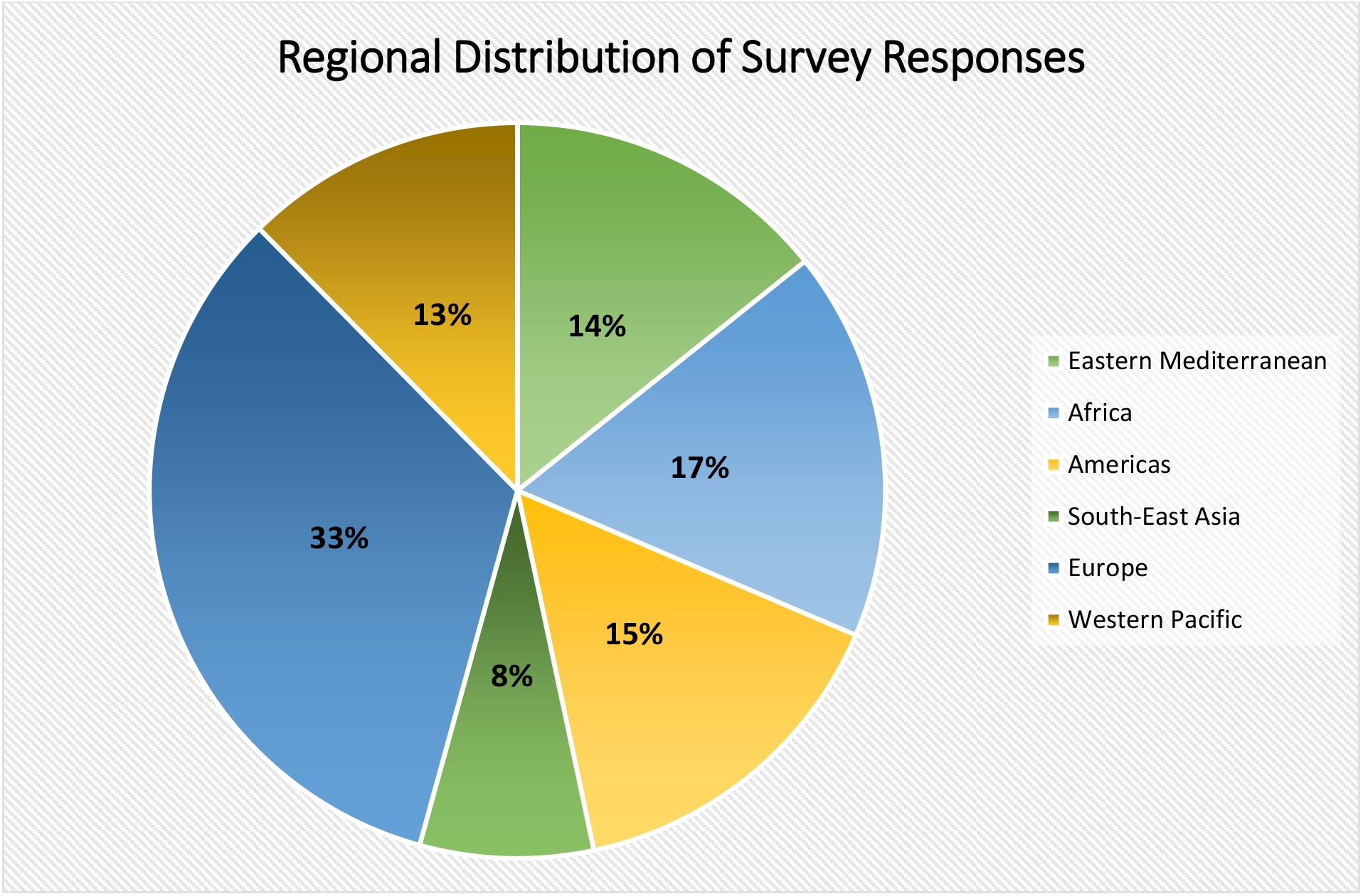
Regional distribution of survey responses.

### NFP Location and Staffing

The overwhelming majority (79%) of States Parties participating in the survey locate their NFP within their Ministry of Health. The remainder place them in closely affiliated ministries, such as the Ministry of Social Affairs, or in national institutes or centres of health or disease control.

59% of the respondents have worked at their NFP office for five years of more; two-thirds have worked there for over eight years (Figure 2). By contrast, nearly 10% of NFPs are relatively new to their work, having been in their NFP office for less than one year.

**Figure 2:**
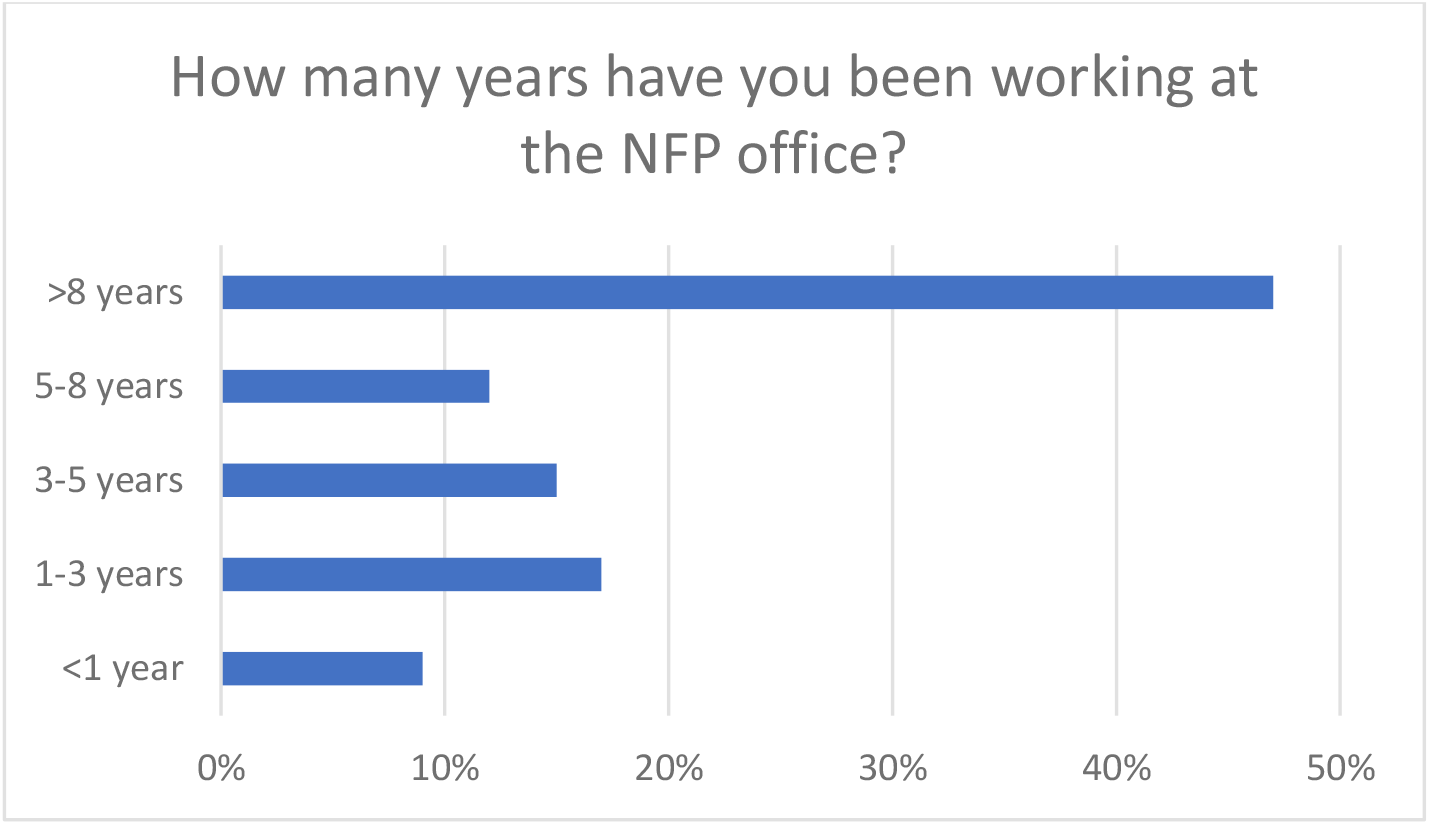
How many years have you been working at the NFP office?

The vast majority (93%), including even those new to their NFP office, believe they are mostly or entirely familiar with the mandatory functions of the NFP under the IHR. NFP staff understood how to contact their designated WHO Regional IHR Contact Point, which NFPS may access at all times for communications concerning the implementation of the Regulations (Article 4.2); only 4% reported that they did not. Similarly, 96% of NFPs reported that they have the necessary content expertise to discuss a notifiable event with the WHO Regional Contact Point.

### Intersectoral collaboration

Only 55% of respondents believe most or all of their colleagues in other ministries and agencies with responsibilities relevant to the implementation of the IHR know how and when to engage with NFPs (Figure 3).

**Figure 3:**
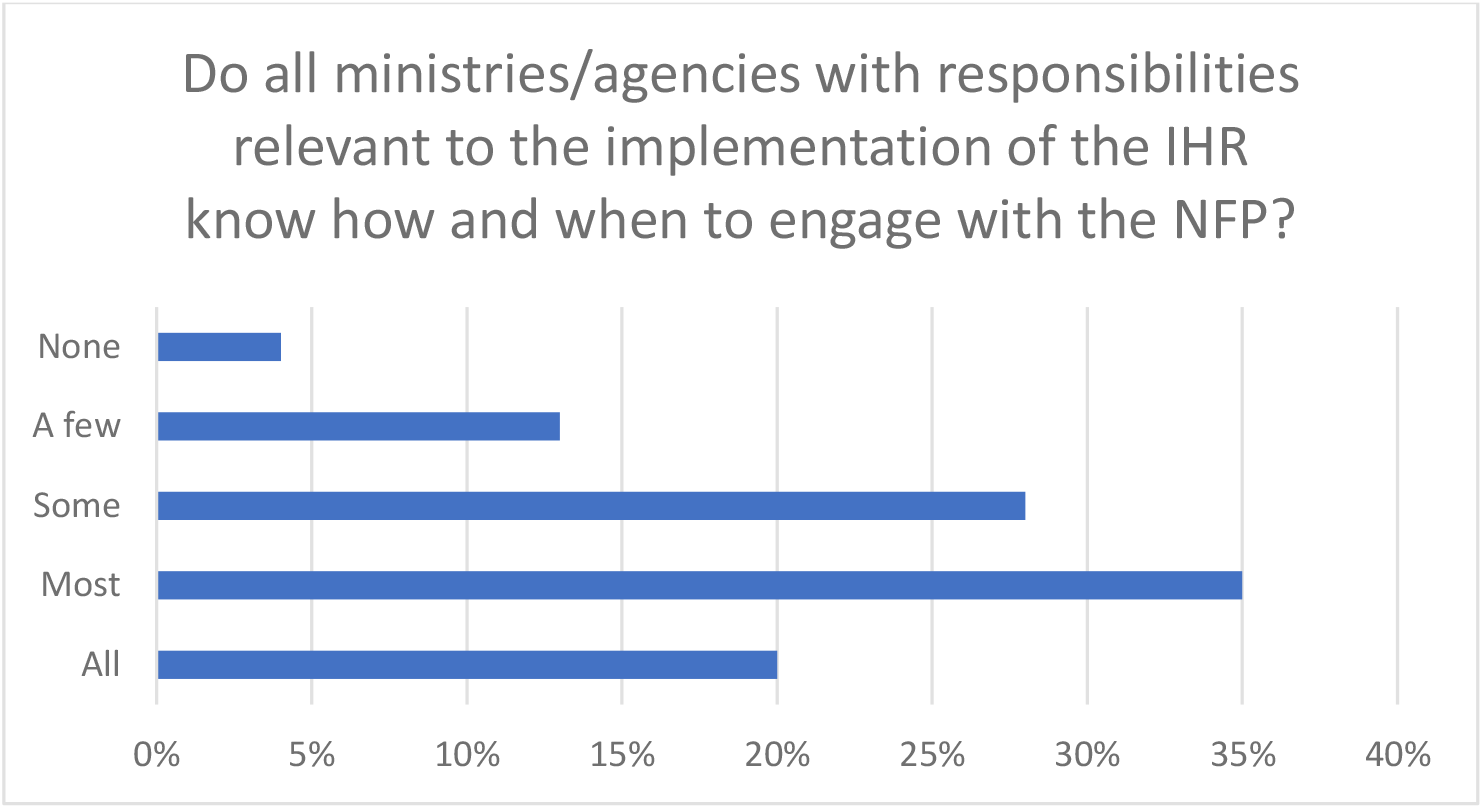
Do all ministries/agencies with responsibilities relevant to the implementation of the IHR know how and when to engage with the NFP?

The questionnaire asked participants whether procedures and structures were in place for timely communication between the NFP and stakeholders in the relevant ministries and agencies. More than 10% did not believe these procedures and structures are in place. Nearly 10% of respondents indicated that their NFPs did not have access to the necessary ministries and decision-makers, including senior management. Over 35% reported that officials in other ministries and agencies must clear NFP notifications to WHO. 15% have never reported an event and did not indicate whether they needed such clearance.

### Communications

The majority of respondents (79%) strongly believe that their NFPs have the ability to send urgent event-related communications to the WHO, whereas 17% only somewhat believe that their NFPs have this ability. Nearly 75% of respondents feel strongly that their NFPs are able to disseminate information from WHO to relevant domestic sectors and consolidate input from these sectors in a timely fashion, while the remainder feel they are only ‘somewhat’ able to do so.

Over 10% of respondents reported that their country does not have a system in place to ensure the NFP is accessible at all times for urgent communications with WHO. Furthermore, about 17% indicated that NFPs do not have adequate information technology systems in place to carry out their IHR communication functions.

Nearly 75% of respondents reported that NFPs sometimes or often communicate bilaterally with NFPs in other countries for assistance or guidance in carrying out their functions or to exchange information (Figure 4).

**Figure 4:**
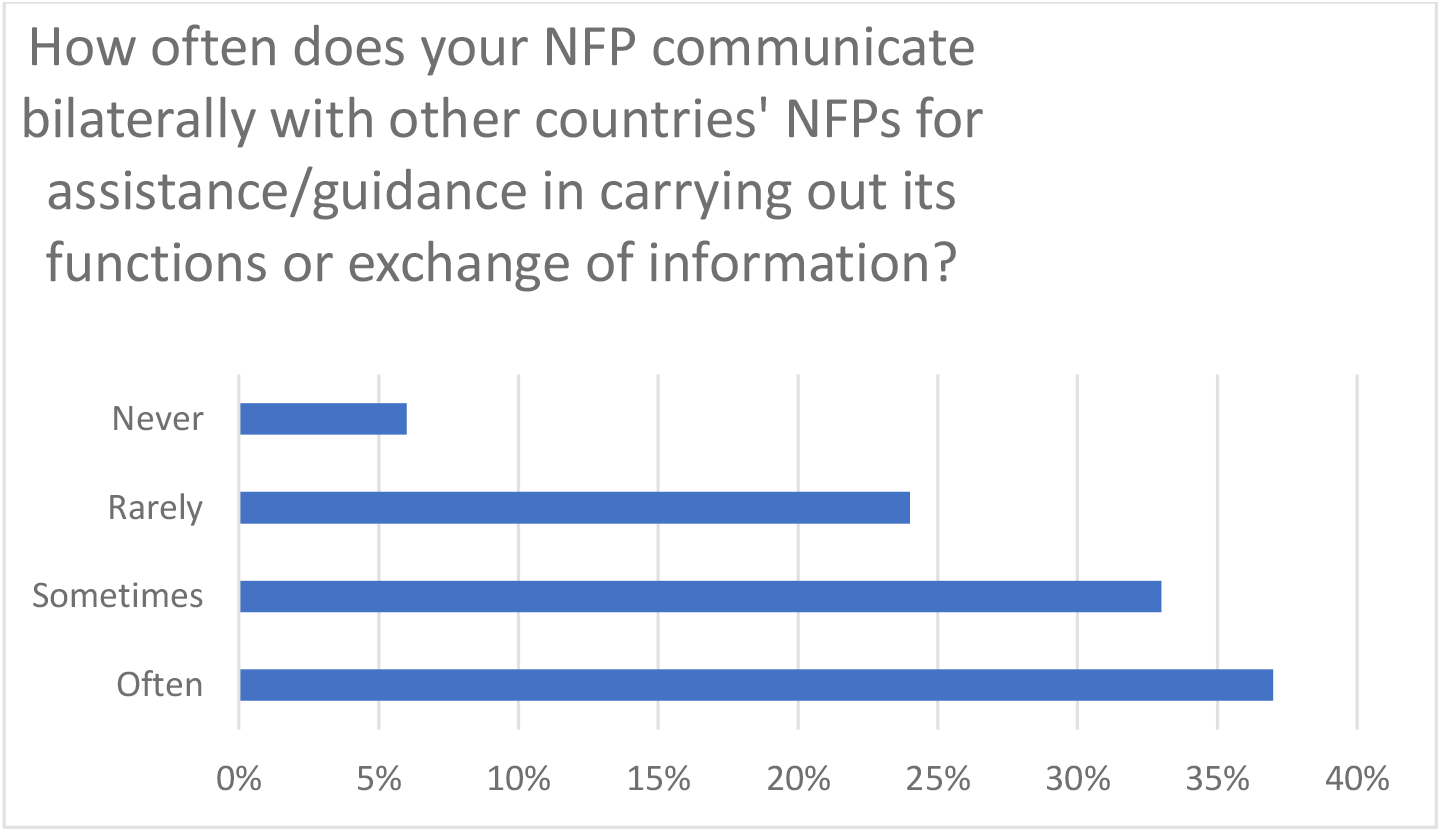
How often does your NFO communicate bilaterally with other countries’ NFPs for assistance/guidance in carrying out its functions or exchange of information?

### Reporting of public health events

13% of survey respondents reported having never submitted a notification of a potential PHEIC to WHO. Of those who had reported such an event, the majority (76%) ‘mostly’ or ‘completely’ based their decision to notify WHO based on the criteria the IHR Annex 2 decision instrument, a tool that helps States Parties decide whether a health event may constitute an urgent public health emergency and requires notification of WHO (8). The remaining 12% who only ‘somewhat’ based their decision to notify WHO based on the criteria set out in Annex 2 (6%), were unsure (2%), or did not use the Annex at all (4%).

The questionnaire asked participants whether they had ever decided not to notify WHO of a potential PHEIC. Only 5% replied positively and reasons included :”it was a local outbreak with no risk of international spread”; “the decision instrument was applied late”; “the [infected] persons had already gone back to their country and were lost to follow up”; “as the secondary designated FP, it is not my responsibility”, and “maybe fear of its impact”.

The questionnaire gave survey participants a list of 11 factors that could impact their timely notification of a potential PHEIC to WHO and asked participants to rate how important these factors are. Figure 5 lists the factors reported, from least to most important.

**Figure 5:**
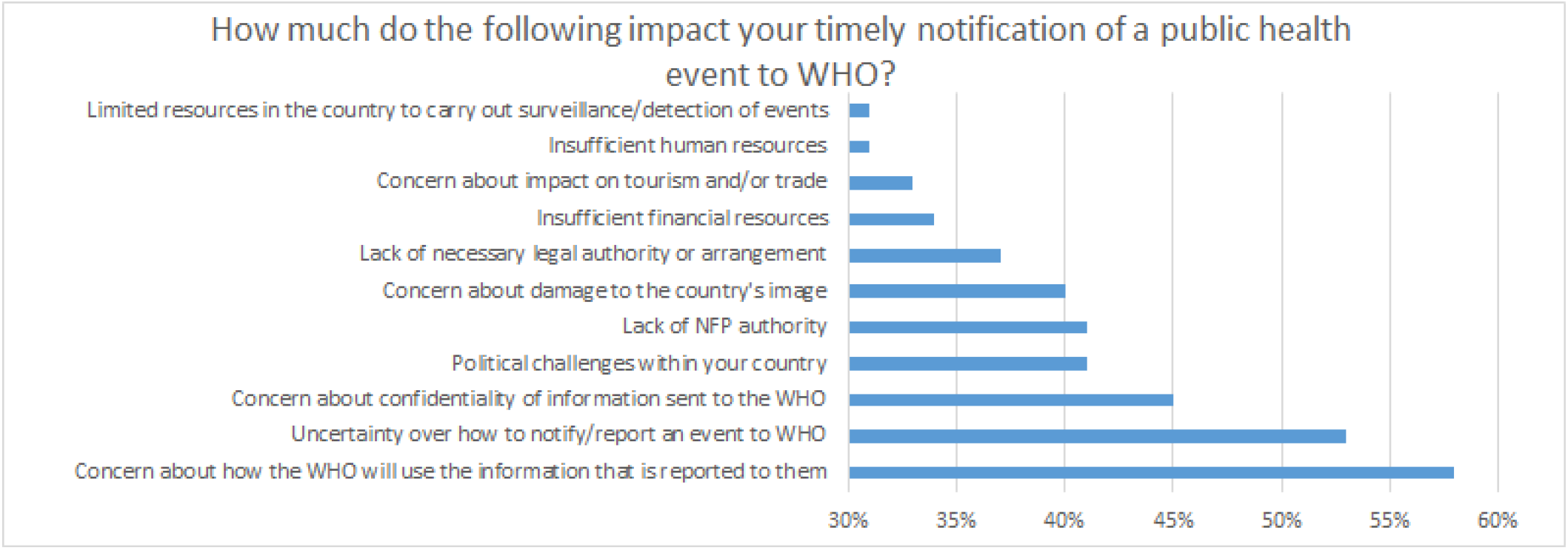
How much do the following impact your timely notification of a public health event to WHO?

The results show that 53% of respondents believe ‘*Concern about how the WHO will use the information that is reported to them* (*e*.*g. dissemination to other countries*)’ is most likely to have impacted their NFPs timely notification of a potential PHIEC to WHO. ‘*Uncertainty over how to report an event*’ followed at 53%.

States Parties were concerned about the potential damage that notification of a possible PHEIC might inflict on their public image (40%) and their tourism or trade (33%). States Parties were also concerned about the confidentiality of reports to WHO (45%). Other factors potentially impacting timely notifications were internal issues such as the NFPs’ lack of the authority to notify WHO (41%), political challenges (41%), and insufficient financial (34%), human (31%) and surveillance/detection (31%) resources.

The survey also asked respondents to rate the adequacy of support they received from different domestic stakeholders and WHO in responding to an event, based on the last incident they had reported to WHO. Of respondents who had submitted at least one notification under the IHR, 70% felt they had received ‘mostly’ or ‘completely’ adequate support from WHO, while the remainder were either unsure or reported receiving only somewhat adequate or inadequate support. A similar number reported adequate support from their national governments, 72% being ‘mostly’ or ‘completely’ satisfied with such support. 24% reported they had received inadequate support, were unsure, or had only somewhat received sufficient support from their national government.

The majority (69%) of respondents reported that their NFPs usually submit notifications to WHO within 24 to 48 hours of determination that an event is notifiable under the IHR. 28% required 48 to72 hours.

The survey asked participants to identify and describe what factors have affected their ability to identify and report notifiable events. Those who offered responses pointed towards the importance of training opportunities and regional networking among of NFPs:

- ‘simulation exercises are a real chance to identify gaps and set evidence-based interventions’;
- ‘with frequent turn-over of staff, more frequent training and orientation on tools should be done’;
- ‘the regional knowledge network is very important to improve our experiences’; and,
- ‘regional NFP meetings organized by the WHO were very useful for sharing experiences’.

The survey asked respondents to describe what more WHO could do to make NFP training more effective in improving NFP ability to identify and report notifiable events. Many responses centred again on capacity-building, with a number of requests for more simulation exercises, training modules for NFPs and other IHR stakeholders, peer-to-peer training through WHO Regional Offices, and more WHO-facilitated opportunities for NFPs to share experiences [and best practices].

Finally, the survey asked participants what mobile or information technology is needed to help NFPs improve their ability to implement the IHR. 38% replied that there was no role for these technologies. Those who identified a need for technological improvement provided the following examples:

- mobile applications to help NFPs disseminate information and reports to stakeholders;
- a dedicated video conferencing system for WHO consultations;
- a hotline for advice;
- a secure reporting portal;
- an interactive platform to assist with IHR interpretation, especially concerning Annex 2; and
- improvements to existing electronic systems (e.g. Early Warning and Response System (EWRS) and Epidemic Intelligence Information System (EPIS)).

### WHO Resources for NFPs

For the majority of NFPs, the WHO-provided NFP Guide (9) is effective (62% declare it to be ‘very useful’). However, 30% believe the Guide to be only ‘somewhat useful’, ‘not very useful’ or ‘not at all useful’. The remainder declared they were not aware of the Guide.

The survey asked respondents to indicate their familiarity with the optional NFP functions outlined in the Appendix of the NFP Guide. 66% of respondents reported that they are ‘mostly’ or ‘completely’ familiar with these, but the remaining believed they are only ‘somewhat’ or ‘not at all’ familiar with them or ‘unsure’.

Respondents largely found the Annex 2 tutorials to be useful (Figure 6).

**Figure 6:**
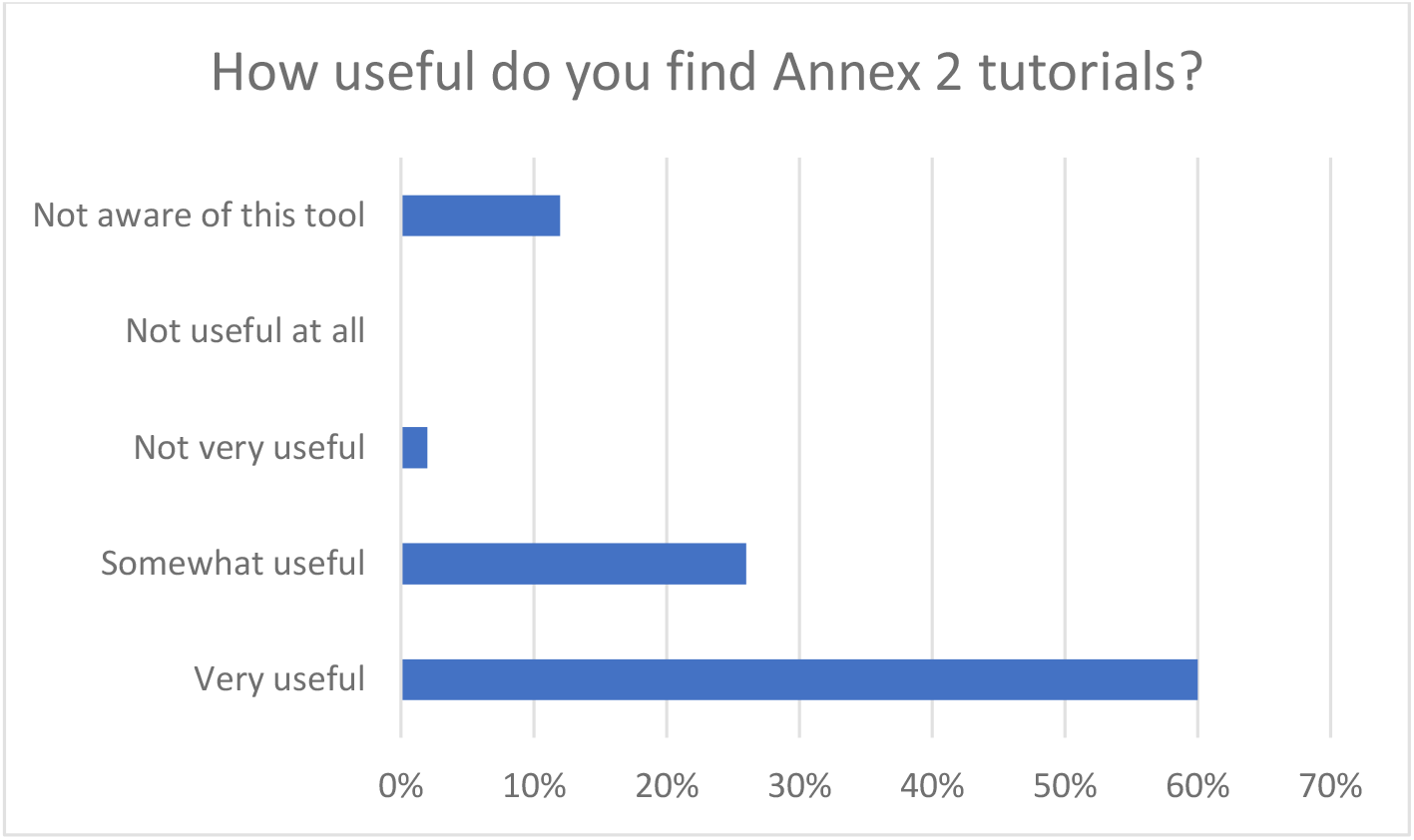
How useful do you find Annex 2 tutorials?

WHO also provides NFPs a Toolkit for Implementation in National Legislation. Responses show that, of all tools provided by WHO, this toolkit was among the least known to NFPs. 23% of respondents did not know it existed and the majority who used the toolkit found it was only somewhat, not very or not at all useful (Figure 7).

**Figure 7.**
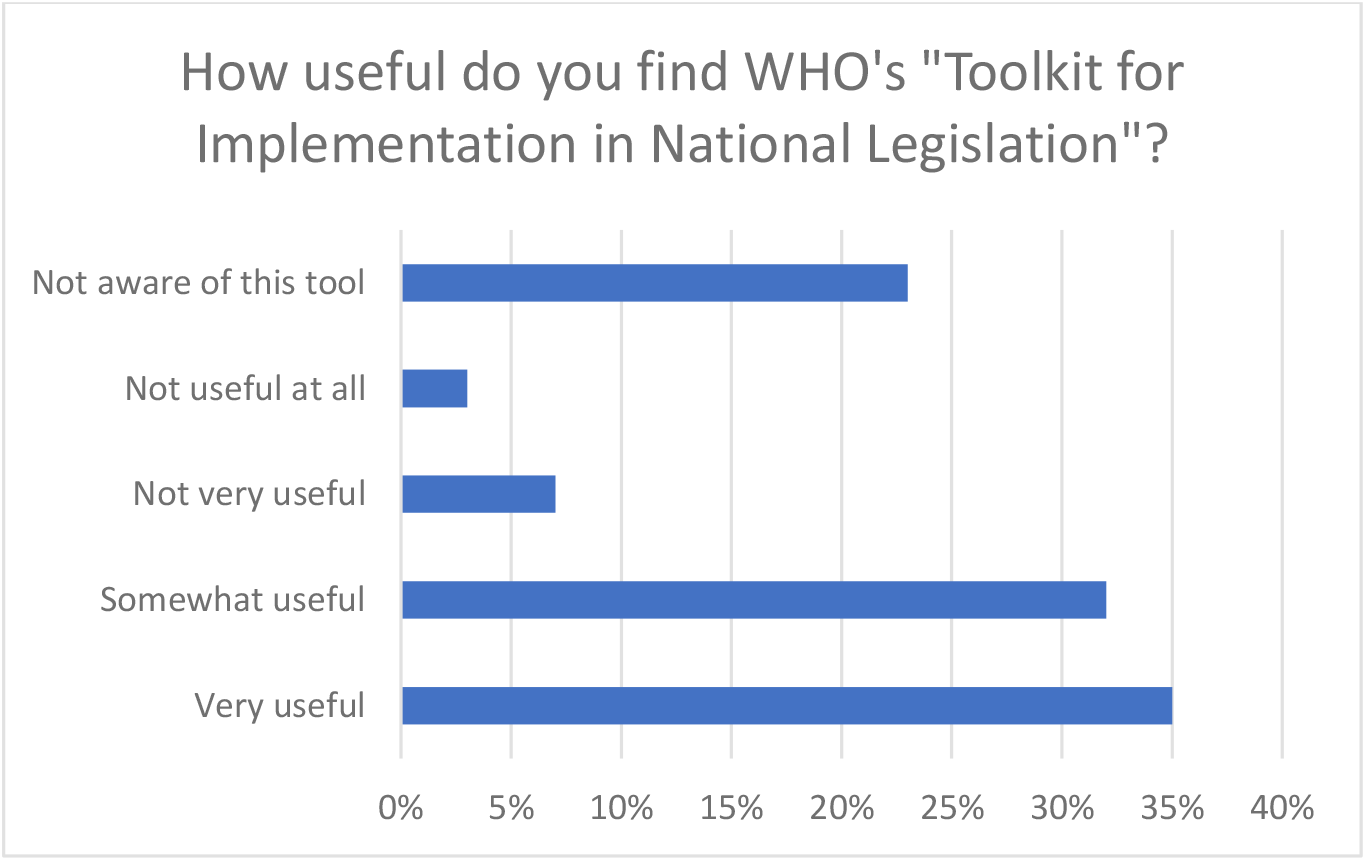
How useful do you find WHO’s “Toolkit for Implementation in National Legislation”?

Respondents were fairly evenly split in rating WHO’s online IHR training course as ‘very’ or ‘somewhat’ useful. 19% of them were not aware this online course existed.

WHO also operates global or regional knowledge networks and organizes workshops by WHO Regional Offices. Of all the WHO resources made available to NFPs, these knowledge networks were the least known; 24% reported not being aware of them. 70% found these networks to be ‘very’ or ‘somewhat’ useful (Figure 8).

**Figure 8:**
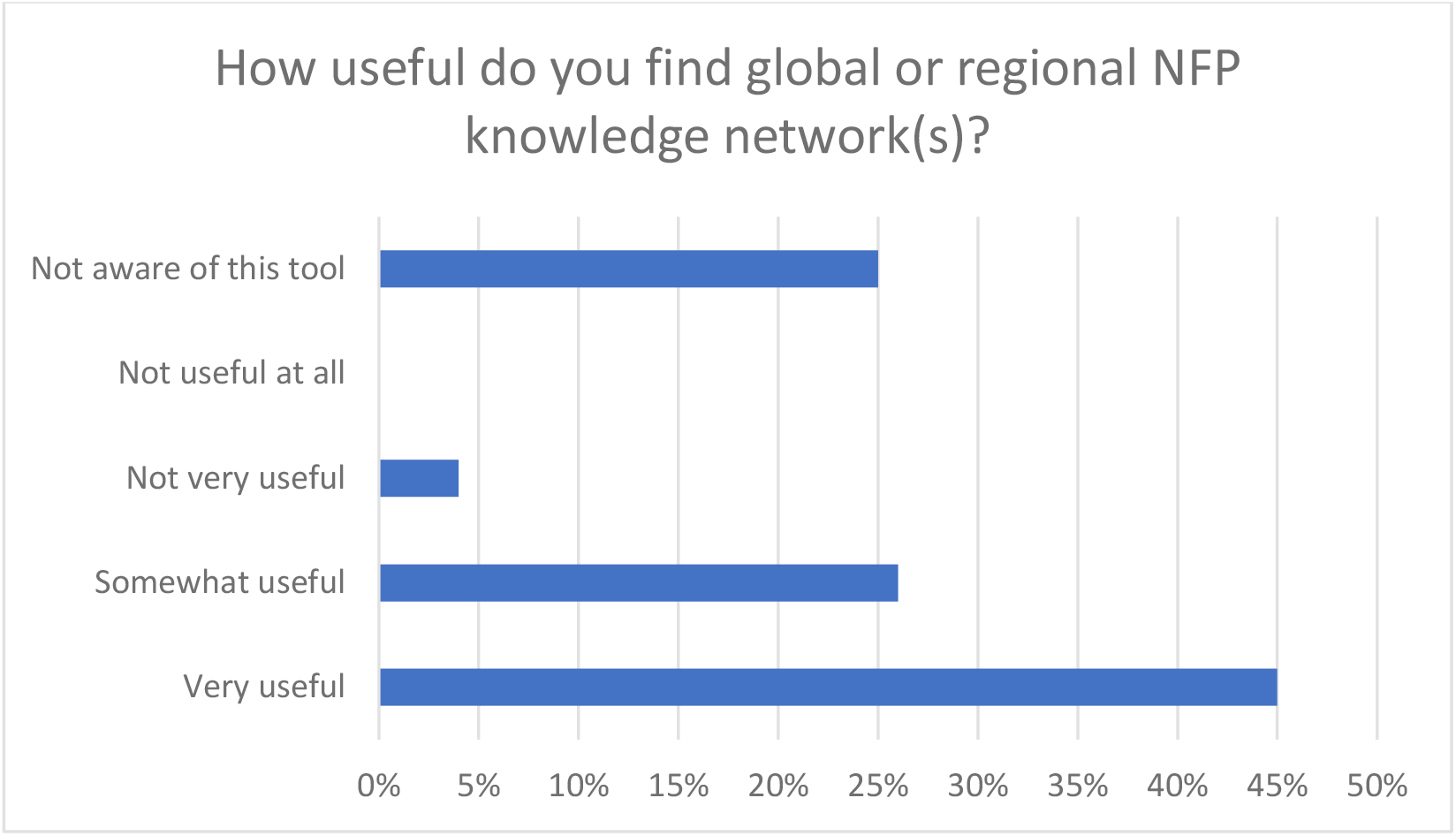
How useful do you find global or regional NFP knowledge network(s)?

NFPs found the workshops organized by WHO Regional Offices the most useful of all WHO resources offered to them. 85% described them as ‘very’ or ‘somewhat’ useful.

Finally, the survey asked participants if they have support the development and learning of staff working in the NFP. The responses were evenly split – 50% did, and 50% did not.

## Discussion

Our study provides an overview of the status, experiences and barriers to effective performance of NFPs prior to the COVID-19 pandemic. We identified positive features and good practices that help NFPs carry out their functions, including sound knowledge of the IHR and reporting procedures, good contact with WHO Regional Contact Points and some useful tools and resources. However, the survey identified challenges related to intersectoral collaboration, communication with WHO and other NFPs, reporting of notifiable events, and WHO resources for NFPs.

The survey results indicate that NFPs are secure in their knowledge and understanding of the IHR, the tasks expected of them, and how to communicate with WHO. However, NFPs appear less confident about whether key colleagues in other ministries understand the IHR and about how such colleagues collaborate on IHR implementation. In order to carry out its functions and interact with WHO in a timely fashion, the NFP depends on input from other ministries and agencies in sectors outside of health. The survey results demonstrate that respondents believe a fair number of colleagues in those sectors have an insufficient understanding of the role of NFPs or know how and when to engage with NFPs. This challenge is not new (10). Some NFPs also lack the procedures and structures to ensure timely communication between the NFPs and national stakeholders in other sectors to support IHR implementation. This challenge is also not new (11). Ineffective intersectoral collaboration can jeopardize the timeliness of information shared and the expeditious reporting of notifiable events to WHO.

NFPs generally know whom to contact at the WHO regional level and how to send urgent event-related communications to WHO, but over a quarter of NFPs reported receiving inadequate support from WHO to help respond to notified events. The survey identified weaknesses in communications that adversely affect NFP implementation of the IHR. For example, some NFPs are not accessible at all times for urgent communications with WHO. Other NFPs do not have the appropriate information technology to carry out NFP communication functions. By contrast, the survey found that communication between NFPs in different States Parties was robust. NFPs are favourable to strengthening this even further if a peer-to-peer learning network was developed and overseen by WHO. They are also favourable towards making regional workshops more available.

It is surprising and worrying that 12% on NFPs report are unsure, only somewhat use or have never used the criteria in Annex 2 to help them decide whether an event must be reported. It is unclear from the fixed responses whether these respondents believe they did not use the right criteria, or whether they never felt an event occurred that merited assessment using the criteria in the Annex 2 tool.

A number of NFPs cannot issue a notification to the WHO without receiving clearance from decision-makers in other domestic sectors owing to lack of legal authority. This matter was raised nearly a decade ago so it is clearly an issue which has been difficult for States to resolve (12). Furthermore, various national political and economic concerns impede NFP implementation of the IHR, including concerns about the confidentiality of information provided to WHO, how WHO would use the information, and the potential damage notifying a potential PHEIC might inflict on a States Party’s image, tourism or trade.

WHO offers resources to help guide and train NFPs, including the NFP Guide, Annex 2 tutorials, the ‘Toolkit for implementation in National Legislation’, an IHR training toolkit course available online, and access to knowledge networks and regional workshops. The survey’s findings on these resources paint a mixed picture. Although the majority of respondents reported they find the resources to be very useful, a significant number find them to be only somewhat, not very or not at all useful, suggesting that improvements can be made. Equally worrying is the number of respondents who are not aware these tools exist. Further, half of those responding report that neither their NFP nor government has a plan to support NFP staff development and learning. NFPs are not sufficiently aware of WHO knowledge networks and workshops. Those NFPs that have used these resources have found them very useful, suggesting that WHO should work to make them more known and available to NFPs.

This study offers some insight into what is and is not working for the NFPs in fulfilling their functions under the IHR. Some problems are easier to address, such as creating more and updated online simulation exercises. Other challenges, such as improving IHR collaboration and communication between national ministries and agencies and addressing the lack of NFP authority to report notifiable events without political clearance, are more difficult to tackle.

Strengths of this study include our ability to access a broad number of NFP’s facilitated by the WHO and their regional offices. Collaboration with the WHO assisted us in developing questions of direct relevance to informing decision-making. The survey questions were also informed by qualitative interviews. Limitations of the study include the response rate. Responders may have been systematically different from non-responders. There is also a risk of social desirability bias with respondents potentially providing responses that they believe they should rather than reflecting the actual situation. While we ensured anonymity, it is plausible this bias still persisted.

These findings can be informative as the current review of the IHR is being conducted and are consistent with their initial recommendations. On 19 January 2021, a summary statement to the WHO was presented to the 148th Executive Board by the Chair of the Review Committee on the Functioning of the IHR (2005) during the COVID-19 Response. It stated that: “National IHR Focal Points need to be further empowered, including where necessary through national legislation. National Focal Points play a critical role in the timely sharing of information, but their limited authority and status often lead to delays in notification. The Committee noted that effective IHR implementation requires many functions that are not within the narrow mandate of the national IHR focal points, such as multisectoral coordination for preparedness and response and collaborative risk assessment(13).”

Examination of the roles of the NFPs at the early stages of the pandemic on providing timely public health information to the WHO and how they could be better supported in this regard are emerging as an important theme. These findings will be relevant as we emerge from the COVID-19 pandemic.

## Data Availability

Data collected is confidential.

## Declarations

### Ethics approval and consent to participate

University of Ottawa REB: H-03-19-3489 - ANN2-348

OHSN REB: 20180438-01H

### Consent for publication

Not applicable

### Availability of data and materials

Not applicable

### Authors’ contributions

CP, SM, LW, KW, HH, AR participated in collecting and analyzing data.

CP, SM, KW wrote the manuscript.

SH, RL, DF, LOG provided critical edits.

All authors read and approved the final manuscript.

## References

1. World Health Organization. International Health Regulations (2005) Third Edition [Internet]. 2016. [cited 2020 Nov 10]. Available from: https://www.who.int/publications/i/item/9789241580496

2. Anema A, Druyts E, Hollmeyer HG, Hardiman MC, Wilson K. Descriptive review and evaluation of the functioning of the International Health Regulations (IHR) Annex 2. Vol. 8, Globalization and Health. 2012.

3. Hardiman MC. World health organization perspective on implementation of international health regulations. Emerg Infect Dis. 2012 Jul;18(7):1041–6.

4. World Health Organization. Implementation of the International Health Regulations (2005) Report of the Review Committee on Second Extensions for Establishing National Public Health Capacities and on IHR Implementation [Internet]. [cited 2020 May 26]. Available from: https://apps.who.int/gb/ebwha/pdf_files/WHA68/A68_22Add1-en.pdf?ua=1

5. World Health Organization. Implementation of the International Health Regulations (2005) Report of the Review Committee on Second Extensions for Establishing National Public Health Capacities and on IHR Implementation Report by the Director-General [Internet]. 2015 Jan [cited 2021 Jan 22]. Available from: https://apps.who.int/iris/bitstream/handle/10665/251717/B136_22Add1-en.pdf?sequence=1&isAllowed=y

6. World Health Organization. Implementation of the International Health Regulations (2005) Report of the Review Committee on the Functioning of the International Health Regulations (2005) in relation to Pandemic (H1N1) 2009 Report by the Director-General [Internet]. 2011 May [cited 2021 Jan 22]. Available from: https://apps.who.int/gb/ebwha/pdf_files/WHA64/A64_10-en.pdf

7. Haustein T, Hollmeyer H, Hardiman M, Harbarth S, Pittet D. WHO | Should this event be notified to the World Health Organization? Reliability of the International Health Regulations notification assessment process [Internet]. 2011 [cited 2020 May 26]. Available from: https://www.who.int/bulletin/volumes/89/4/10-083154/en/

8. World Health Organization. WHO Guidance for the Use of Annex 2 of the INTERNATIONAL HEALTH REGULATIONS (2005) [Internet]. 2008 [cited 2020 Nov 10]. Available from: https://www.who.int/ihr/revised_annex2_guidance.pdf?ua=1

9. World Health Organization. Designation/establishment of National IHR Focal Points [Internet]. 2006 Jul [cited 2021 Jan 22]. Available from: https://www.who.int/publications/m/item/designation-establishment-of-national-ihr-focal-points

10. Wamala JF, Okot C, Makumbi I, Natseri N, Kisakye A, Nanyunja M, et al. Assessment of core capacities for the International Health Regulations (IHR[2005]) - Uganda, 2009. BMC Public Health. 2010 Dec 3;10(SUPPL. 1):S9.

11. Suthar AB, Allen LG, Cifuentes S, Dye C, Nagata JM. Systematic reviews Lessons learnt from implementation of the International Health Regulations: a systematic review. Bull World Heal Organ. 2018;96:110–21.

12. Burkle FM, Redmond AD, McArdle DF. An authority for crisis coordination and accountability. Lancet. 2012 Jun 16;379(9833):2223–5.

13. World Health Organization. Statement to the 148th Executive Board by the Chair of the Review Committee on the Functioning of the International Health Regulations (2005) during the COVID-19 Response [Internet]. 2021 [cited 2021 Jan 22]. Available from: https://www.who.int/news/item/19-01-2021-statement-to-the-148th-executive-board-by-the-chair-of-the-review-committee-on-the-functioning-of-the-international-health-regulations-(2005)-during-the-covid-19-response

